# SARS-CoV-2 reinfections with BA.1 (Omicron) variant among fully vaccinated individuals in the northeast of Brazil

**DOI:** 10.1101/2022.04.08.22272726

**Authors:** Francisco P. F. Neto, Diego G. Teixeira, Dayse C. S. da Cunha, Ingryd C. Morais, Celisa P. M. Tavares, Genilson P. Gurgel, Sanderson D. do Nascimento, David C. dos Santos, Alexandre de O. Sales, Selma M.B. Jerônimo

## Abstract

**Background:** The first case of Severe Acute Respiratory Syndrome Coronavirus 2 (SARS-CoV-2) in Rio Grande do Norte, northeast Brazil, was diagnosed on March 12, 2020; thereafter, the pattern of COVID-19 followed the multiple waves as seen elsewhere. Those waves were mostly due to the SARS-CoV-2 virus mutations leading to emergence of variants of concern (VoC). The introduction of new VoCs in a population context of prior SARS-CoV-2 infections or after vaccination has been a challenge in understanding the kinetics of the protective immune response against SARS-CoV-2. The aim of this study was to investigate the outbreak of SARS-CoV-2 reinfections observed in mid-January 2022 in Rio Grande do Norte state, Brazil when the omicron variant was introduced.

**Methodology/Principal findings:** From a total of 172,965 individuals with mild to severe respiratory symptoms, 58,097 tested positive for SARS-CoV-2 between March 2020 through mid-February 2022. Of those previously infected, 444 had documented a second SARS-CoV-2 infection and 9 of these reinfection cases were selected for sequencing. Genomic analysis revealed that virus lineages diverged between primary and the reinfection, with the latter caused by the Omicron (BA.1) variant among individuals fully vaccinated against SARS-CoV-2.

**Conclusions/Significance:** Once all subjects whose samples were sequenced had prior SARS-CoV-2 infection and were also fully vaccinated, our data suggest that the Omicron variant evades natural and vaccine-induced immunities, confirming the continuous need to decrease transmission and to develop effective blocking vaccines.

**Author summary:** The pattern of the COVID-19 pandemic has been characterized by multiple waves of cases with a variety of outcomes from asymptomatic, to moderate or to severe fatal cases. By December 2021, about 75.3% of Rio Grande do Norte population, northeast Brazil, had already been fully vaccinated against SARS-CoV-2 and a decrease in newer detection cases was seen to about 8% of the suspected ones. Nevertheless, with the introduction of the Omicron variant at the end of 2021, the number of new SARS-CoV-2 infections reached its highest peak since the start of the pandemic with 75% of the suspected cases testing positive. From March 2020 to February 2022, we confirmed 444 reinfection cases among the ones tested, of which 62.3% (n=277) occurred during the Omicron outbreak, from December 2021 to early February 2022. Of the reinfection cases, 9 were sequenced and genetic analysis showed that they belong to a BA.1 lineage, which seems to have been introduced multiple times into the region. The primary isolates varied. Thus, our data suggest that the Omicron variant evades immunity provided from either natural infection from any other SARS-CoV-2 variants or from different types of vaccines.

## Introduction

The world has followed the spread of SARS-CoV-2 since 2019 in real time. However, over two years after the first cases of Coronavirus Disease 2019 (COVID-19) were reported in China, much is still unknown about the mutations and the molecular evolution of SARS-CoV-2 over time. The unpredictability of host diversity and the selective pressure by the immune system either by natural infection and/or vaccination on viruses lead us to a field of concerns that gradually have been enlightened.

The emergence of SARS-CoV-2 variants with potentially increased transmissibility, virulence, and resistance to antibody neutralization have already been reported [1,2]. The dissemination of these variants, so-called variants of concern (VoC), are also closely associated with higher viral load [3], and/or rates of reinfection [4] among individuals. By the fact that VoCs can surpass the protective effects of humoral immunity induced either by natural infection or by vaccination [5,6], this poses an additional challenge to curb the pandemic since the highest number of new cases and an increased number of reinfections still have been reported worldwide in 2022 [7].

The multiwave pandemic dynamics plays different scenarios depending on the mutations present in each SARS-CoV-2 variant and its location. Some of them can provide great impact in specific regions of the globe. For example, studies have shown that the Gamma (P.1) variant is more prevalent in South America [8–10], however, by contrast, the Delta variant and its sublineages [4,11,12] have been found all over the world.

Recently, Brazil faced the third wave of COVID-19 pandemic with a record upsurge of cases that started in the end of December 2021 and it had been sustained by the highly transmissible variant B.1.1.529, a VoC, named Omicron [13]. In Brazil, COVID-19 vaccination started on January 17, 2021, however despite the high coverage of immunization in the country, achieving over 79% of the population with at least one dose of the vaccine, at the end of January 2022 [14], there was still a sustained number of cases occurring over the year of 2021, and some of the COVID-19 reinfection cases had been reported among fully vaccinated people [6]. From March 2020 to February 2022, 444 reinfection episodes were detected in our database, in which 277 cases were diagnosed in the first two months of 2022. Here, we present nine cases of reinfection with the Omicron variant in immunocompetent and fully vaccinated individuals. We also provide genomic variations and clinical features between primary infection and reinfection cases among residents of Rio Grande do Norte (RN) State, located in the Northeastern region of Brazil.

## Materials and Methods

### Data acquisition, sample collection and clinical testing

All clinical data and nasopharyngeal swab specimens from the first and second infection were obtained from patients in the municipalities of Natal and Apodi, located in the state of Rio Grande do Norte, Brazil. The research was carried out at the Instituto de Medicina Tropical-UFRN (IMT-UFRN) in a partnership with Getúlio Sales Diagnósticos and Health Municipal Secretariats of Apodi and Natal.

Nasopharyngeal swabs were collected and placed in Viral Transport Medium. Samples were transported to IMT-UFRN and tested in the same day for SARS-CoV-2 by reverse transcriptase quantitative polymerase chain reaction (RT-qPCR). Remnant samples were stored at −80 °C. SARS-CoV-2 infection was diagnosed by RT-qPCR using CDC/EUA protocol [15]. The volunteers filled a questionnaire with epidemiological questions We selected 18 samples with RT-qPCR positive results, presenting a cycle threshold (Ct) value ≤ 30, collected from July 2020 to February 2022.

### RNA extraction, library preparation and sequencing

Viral RNA was isolated from 200-μl samples collected from people suspected of SARS-CoV-2-infections using the QIAamp Viral RNA Mini kit (QIAGEN, Hilden, Germany) according to the manufacturer’s instructions. Virus genome sequencing was carried out on all positive samples selected using a COVIDSeq protocol (Illumina Inc, USA).

cDNA synthesis was performed from the extracted RNA using a multiplex polymerase chain reaction (PCR) protocol, producing 98 amplicons across the SARS-CoV-2 genome (https://artic.network/). The primer pool additionally had primers targeting human RNA, producing an additional 11 amplicons. The PCR amplified product was later processed for tagmentation and adapter ligation using IDT for Illumina Indexes. Further enrichment and cleanup were performed as per protocols provided by the manufacturer (Illumina Inc). All samples were processed as batches in a 16-well plate. Pooled samples were quantified using Qubit 2.0 fluorometer (Invitrogen Inc.) and fragment sizes were analyzed in 4150 TapeStation System (Agilent Inc). The pooled library was further normalized to 4nM concentration and 25 μl of each normalized pool containing index adapter set A were combined in a new microcentrifuge tube to a final concentration of 100pM. The pooled libraries were loaded and clustered onto iSeq 100 i1 Reagent v2 (Illumina) with flow cell and sequenced using sequencing by synthesis (SBS) chemistry on the Illumina iSeq 100 sequencing system. The sequencing run was created with Local Run Manager software installed on the iSeq 100 instrument. Using the Generate FASTQ Analysis module with the custom library preparation kit options, the number of sequencing cycles was set to 151 bp and the paired-end sequencing option was determined.

### Read mapping and Consensus calling

All reads were mapped to the reference SARS-CoV-2 genome (GenBank accession number MN908947.3) using the BWA-MEM V0.7.12-r1039 [16]. Next, the removal of duplicate reads and the split of reads containing poorly sequenced base was proceeded by the Genomic Analysis-Toolkit (GATK) V4.2.0.0 [17]; the quality of the final mapping process was accessed by Mosdepth [18] and Qualimap V2.2.1 [19]. The SNV identification was performed by the *HaplotypeCaller* function from the GATK, followed by the filtration using the *VariantFiltration, FilterVcf* and *SelectVariants*, excluding variants with depth lower than 10. Those variants which passed by the depth threshold were annotated using the SnpEff V5.0e [20].

The sequence consensus calling was performed by the *FastaAlternateReferenceMaker* function from GATK, and for the final fasta file those positions covered by fewer than 10 reads were replaced by Ns, by using an in-house script written in Python 3.8 (available at GitHub). All sequences were submitted to the Pangolin COVID-19 Lineage Assigner v.3.1.17 [21] (accessed on 24/Jan/2022). The whole genome sequences of viral variants of the nine samples (two samples from each case of reinfection) were deposited in the GISAID database (accession nos. EPI_ISL_9467461, EPI_ISL_9467460, EPI_ISL_9467471, EPI_ISL_9467463, EPI_ISL_9467462, EPI_ISL_9467470, EPI_ISL_9467458, EPI_ISL_9467469, EPI_ISL_9467457, EPI_ISL_9467468, EPI_ISL_9467459, EPI_ISL_9467454, EPI_ISL_9467465, EPI_ISL_9467464, EPI_ISL_9467456, EPI_ISL_9467467, EPI_ISL_9467455, EPI_ISL_9467466) and all GISAID sequences used in this study are listed in S1 Table.

### Phylogenetics analysis

The 18 genome sequences obtained in this work were aligned to 700 other genome sequences from Rio Grande do Norte, Brazil, obtained from GISAID on January 20, 2022, using the MAFFT v.7.394 [22]. We then built a maximum likelihood (ML) phylogenetics tree supported by 1,000 replicates of bootstrap using IQ-TREE2 [23], under the substitution model GTR+F+R4, which was inferred to be the best-fitting model by de software itself. The same procedure was performed for the Alignment containing only the Brazilian BA.1 genomes, under the GTR+F+R3 model. Tree topologies were inspected by Figtree v.1.4.4 (http://tree.bio.ed.ac.uk/software/figtree/ last accessed 2021-11-29).

### Ethical Issues

The study protocol was reviewed and approved by the Federal University of Rio Grande do Norte Ethical committee (36287120.2.0000.5537).

## Results

### Tracking reinfection cases in Rio Grande do Norte, Brazil

From March 2020 to February 2022, a total of 172,965 individuals with upper respiratory symptoms underwent testing by RT-qPCR for SARS-CoV-2 at IMT-RN, of which 58,097 tested positive. Positive tests remained high from December 2020 to July 2021 (mean 160.9/day, 100.4 SD), when thereafter a decrease of COVID-19 cases was observed until the beginning of January 2022 (mean 27.8/day, 28.5 SD), followed by a rapid increase in positivity (Fig 1A). Among those, 444 individuals had 2 episodes of symptomatic infection for COVID-19 (Fig 1B). In mid-2020, reinfection cases were already reported, although scarce, and in early 2022, during the Omicron-predominant period, they reached a peak comprising 62.3% (n=277) of all reinfection cases identified in our system. The earliest day of reinfection was 62 days apart, after the primary infection, and the longest was 638.

**Fig 1.**
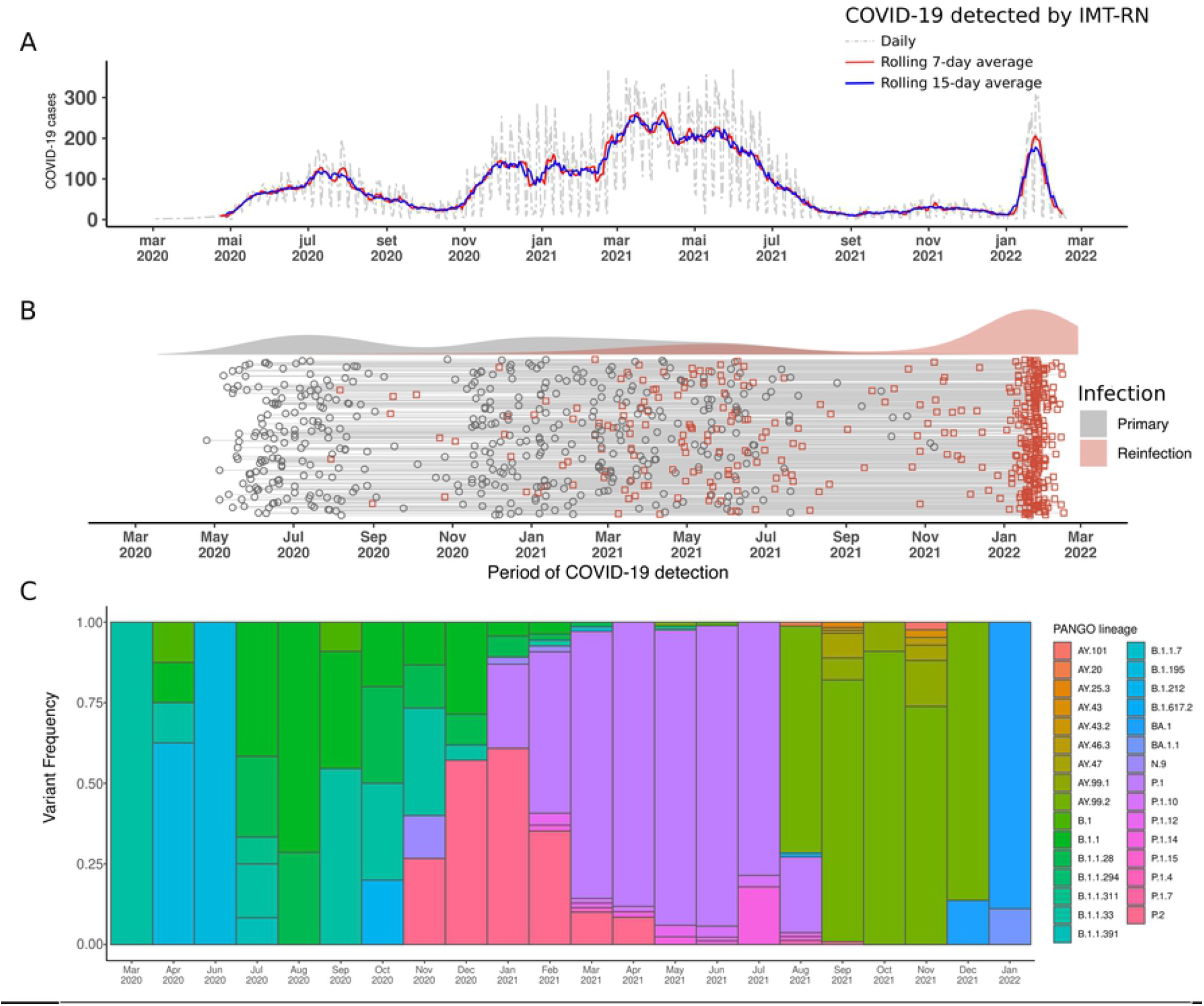
Timeline of COVID-19 detection cases in Rio Grande do Norte state, Brazil. (A) Positive COVID-19 cases in RN detected by RT-qPCR in IMT-RN laboratory. (B) Detection of SARS-CoV-2 infection in 444 individuals. Participants with two entries in our database were selected and the period of the first (gray circle) and second (red square) entry was plotted. A gray line is connecting both COVID-19 tests of the same individual. The density of COVID-19 cases on top of the plot. (C) SARS-CoV-2 PANGO lineages detected in RN from March 2020 to January 2022, obtained from GISAID.

### SARS-CoV-2 isolates sequencing

Of the 444 individuals with documented SARS-CoV-2 reinfection, 9 immunocompetent and fully vaccinated individuals were chosen to have their isolates sequenced (S1 Fig). Among those, 5 were males and the mean age was 32.4 years (± 7.4). The first infection ranged from June 2020 to November 2021. All reinfection cases occurred in January 2022. The time course between the 2 episodes of COVID-19 ranged between 70 and 584 days, with mild clinical presentation for both infections (Fig 2A). The average between the last vaccination date and the second episode was 86.8 days (± 46.1). The most reported symptoms during both infections were headache, nasal congestion, fatigue, rhinorrhea, myalgia, pharyngitis, fever, and cough. Comorbidities, such as hypertension, diabetes mellitus, cancer, and asthma/COPD were not reported among participants and none of them were hospitalized. In addition, two of the subjects had already received a third dose of the vaccine.

**Fig 2.**
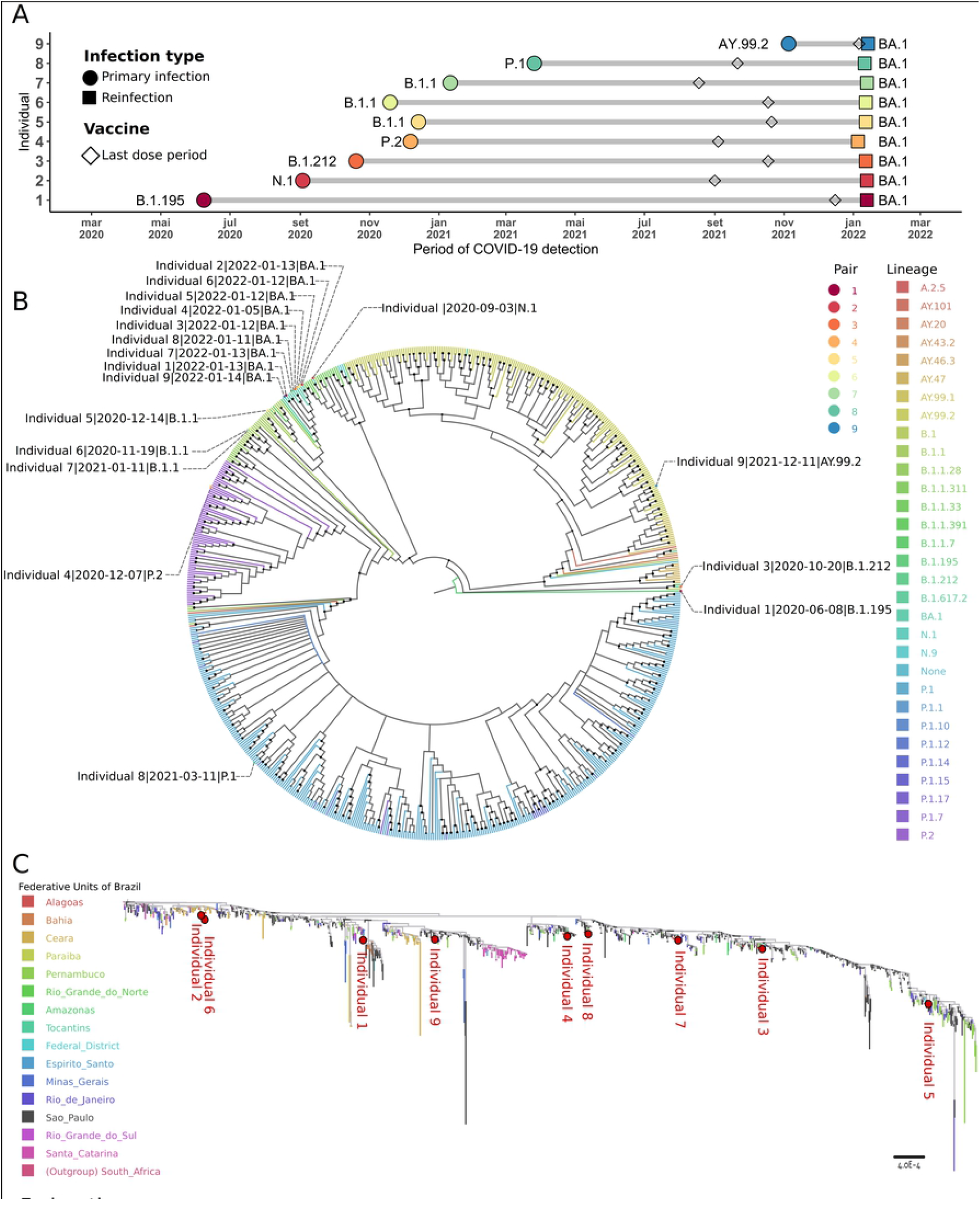
Identification of SARS-CoV-2 reinfection cases in Rio Grande do Norte, Brazil. (**A**) Primary infection and reinfection and the virus lineages. The figure also shows the time of the last COVID-19 vaccine. (**B**) Maximum-likelihood tree of all 709 SARS-CoV-2 whole-genome sequences available at GISAID repository, downloaded on January 20, 2022. The color assigned to the branches are related to their respective lineage and sequenced samples in this work are labeled according to their pair, sampling date and virus lineage, respectively. The dark dots at the dichotomies represent those nodes with high bootstrap support (≥ 80). The tree’s root was set on the youngest isolate (from June 8, 2020), sequenced in this work, which belongs to the B.1.195 lineage. (**C**) ML tree composed by other 1700 Omicron sequences isolated in Brazil plus a B.1.1.529 reference as outgroup (EPI_ISL_6704867), with the branches colored according to the Brazilian state where the virus was isolated. The red dots indicate where each of the 9 sequences are placed in the tree. Each of the 9 isolates and the other sequences from Brazil close to them can be seen in S2 Fig.

In order to distinguish between reinfection and long-term viral persistence, we performed a maximum-likelihood (ML) phylogenetic analysis of all SARS-CoV-2 whole genomes from Rio Grande do Norte state, recovered from GISAID on January 20, 2021, combining with all 18 samples sequenced in this work (Fig 2B). The earliest isolate reported from Rio Grande do Norte state was obtained on July 02, 2020 and belongs to the BA.1.1.311 lineage (EPI_ISL_2466179), but herein we sequenced an oldest one, from June 08, 2020; which was assigned to the B.1.195 lineage by the Pangolin tool, when the ML tree root was set. The tree topology revealed that all sequences belonged to the variant of concern Omicron (BA.1) lineage and they were clustered in a monophyletic clade while their respective pairs, from the previous SARS-CoV-2 infection, were found spread through the tree. A sequence from the individual #2 isolated during the first infection which was assigned to the N.1 lineage, was placed close to the Omicron branch. The N.1 lineage had been detected in Brazil during the early epidemic in 2020, according to the *outbreak*.*info* website.

Among the 9 individuals in this study, a female individual had a reinfection episode 70 days apart from primary infection and 8 days after the COVID-19 vaccine booster shot. Despite this short interval, the virus lineages diverged between the two infections (Figs 2A and B), the primary occurring through a AY.99.2 variant, while the reinfection episode was by BA.1. It is important to note the high frequency of AY lineages (Delta VOC) on the topology ML tree, representing more than 30% of all sequences on Rio Grande do Norte state, and also that variants arising from Delta were found on almost 100% of sequenced isolated in Brazil in December 2021 [24].

To better characterize the Omicron variant detected during the reinfection cases, we aligned it against all other Brazilian BA.1 whole genomes available in the GISAID database until 20 January 2022. In addition, we also selected an African B.1.1.529 reference sequence as an outgroup (accession EPI_ISL_6704867). The second ML tree (Fig 2C) revealed that in a broader scale of this single lineage, the genomes isolated from the reinfection are spread through the tree, emphasizing the multiple introductions of the BA.1 lineage into the Rio Grande do Norte state. The relationship of the 9 BA.1 isolates sequenced here can be observed closer on S2 Fig, with a remarkable presence of isolates from the states of São Paulo, Pernambuco and Ceará.

## Discussion

SARS-CoV-2 infection episodes by emerging variants in previously naturally infected or vaccinated individuals are already a real phenomenon worldwide which poses an important issue to understand human immune response to the virus and vaccines [7]. Reinfection cases have been rising profoundly since the Omicron variant of SARS-CoV-2 started to disseminate in November 2021 [25]. To date, such a fact was not observed with the same rate in previously reported variants of concern of SARS-CoV-2. In our study, we demonstrate that 62.3% (277/444) of all reinfection cases detected over 22 months occurred during the Omicron-wave (Fig 1). To confirm those reinfection events, we proposed to investigate a small prospective cohort study from 9 individuals with reinfection of COVID-19 in the state of Rio Grande do Norte, Brazil. This state receives a high number of tourists during the end of the year, and this may have been one of the reasons for the diversity of introductions in the region. From early-December 2021 to mid-January 2022, São Paulo was the Brazilian state with the most expressive number of flights to Natal (S3 Fig), which may explain the similarities between the majority of the isolates sequenced in this work and the ones sequenced in São Paulo. This, together with the fact that São Paulo is one of the states most affected by the omicron variant [26], due to being a hub for intercontinental flights, provides, therefore, an increased risk of arrival of new variants as the omicron case shows.

The Brazilian Health Ministry criteria for a SARS-CoV-2 reinfection case is two positive RT-qPCR confirmations within at least 90 days apart, clinical manifestations characteristics to COVID-19, positive viral culture, with an additional viral RNA sequencing from both episodes showing different strains [6],[27]. Our work lacks the viral culture, nevertheless, we demonstrated one case of an individual with two infection episodes, 70 days apart, and the sequence of the two isolates showed clearly different lineages. Reinfection cases under 90 days apart from the episodes have already been described [28].

Recent studies have demonstrated that Omicron-infected cases tended to cause mild symptoms such as pharyngitis, headache, nasal [29] which corroborates with our data. Despite the fact that three subjects have a booster, with the third/additional dose, they developed symptomatic COVID-19, as others with complete vaccination and previous history of COVID-19 [7]. These data support the findings that neither the current vaccines available nor previous history of COVID-19 are sufficient to establish a robust response toward the Omicron variant and to promote a reduced risk of infection. Nonetheless, a recent study has shown that booster vaccination increases the neutralization titers to Omicron [5].

Although with the data presented herein, we have barely scratched the surface of the many possibilities in which the Omicron lineage or other variants are capable of causing reinfections, further studies are needed to understand the role of SARS-CoV-2 mutations which lead the evasion of the immune response generated by prior infection or vaccination. The kinetics of the protective immunity is quite not ascertained. The currently available vaccines have apparently reduced severity of the COVID-19, even though an increase in hospitalization and deaths occurred with the introduction of the apparent more benign omicron variant, when compared to the Gamma one. This indicates the need to develop more effective vaccines, with potential for reducing transmission and a lengthier protective immune response. Genomic surveillance is a key for assessing new variants and therefore suitable for planning new strategies to battle COVID-19.

Our data suggest that the Omicron variant evades immunity provided from different types of vaccines or from natural infection from any other earlier SARS-CoV-2 variants of concern and may be related mostly to the mild symptoms of the upper respiratory tract for the majority of the subjects (S1 Fig). Thus, along with vaccination, the reassurance of protective measures needs to be held to prevent further spread of SARS-CoV-2 variants or risk of appearance of new variants.

## Data Availability

All data produced in the present work are contained in the manuscript

## Acknowledgements

We would like to thank the Núcleo de Processamento de Alto Desempenho da Universidade Federal do Rio Grande do Norte - NPAD/UFRN for allowing us to access their computer facilities.

## Supporting Information Captions

**S1 Table**. GISAID sequences used in this study.

## SUPPLEMENTARY RESULTS

**Supplementary Figure S1.**
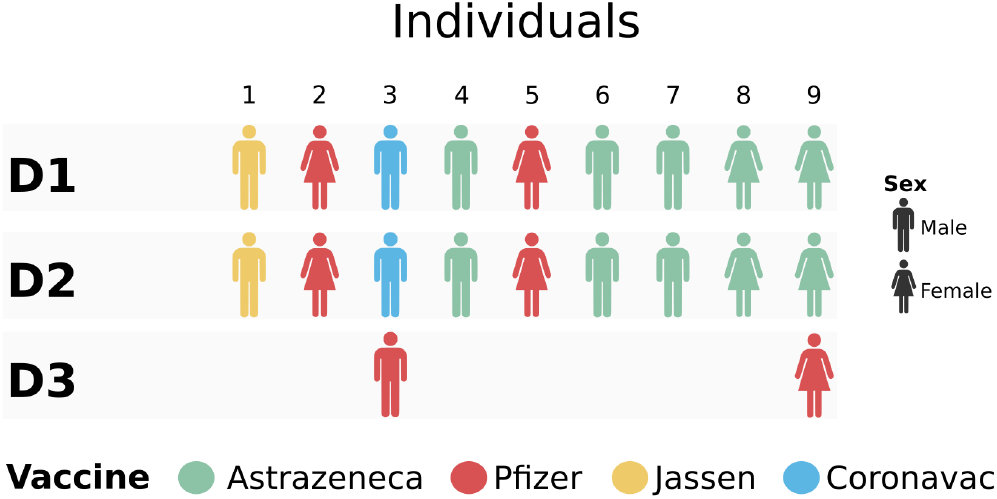
Schematic representation of different types of vaccine against SARS-CoV-2 in all reinfection cases.

**Supplementary Figure S2.**
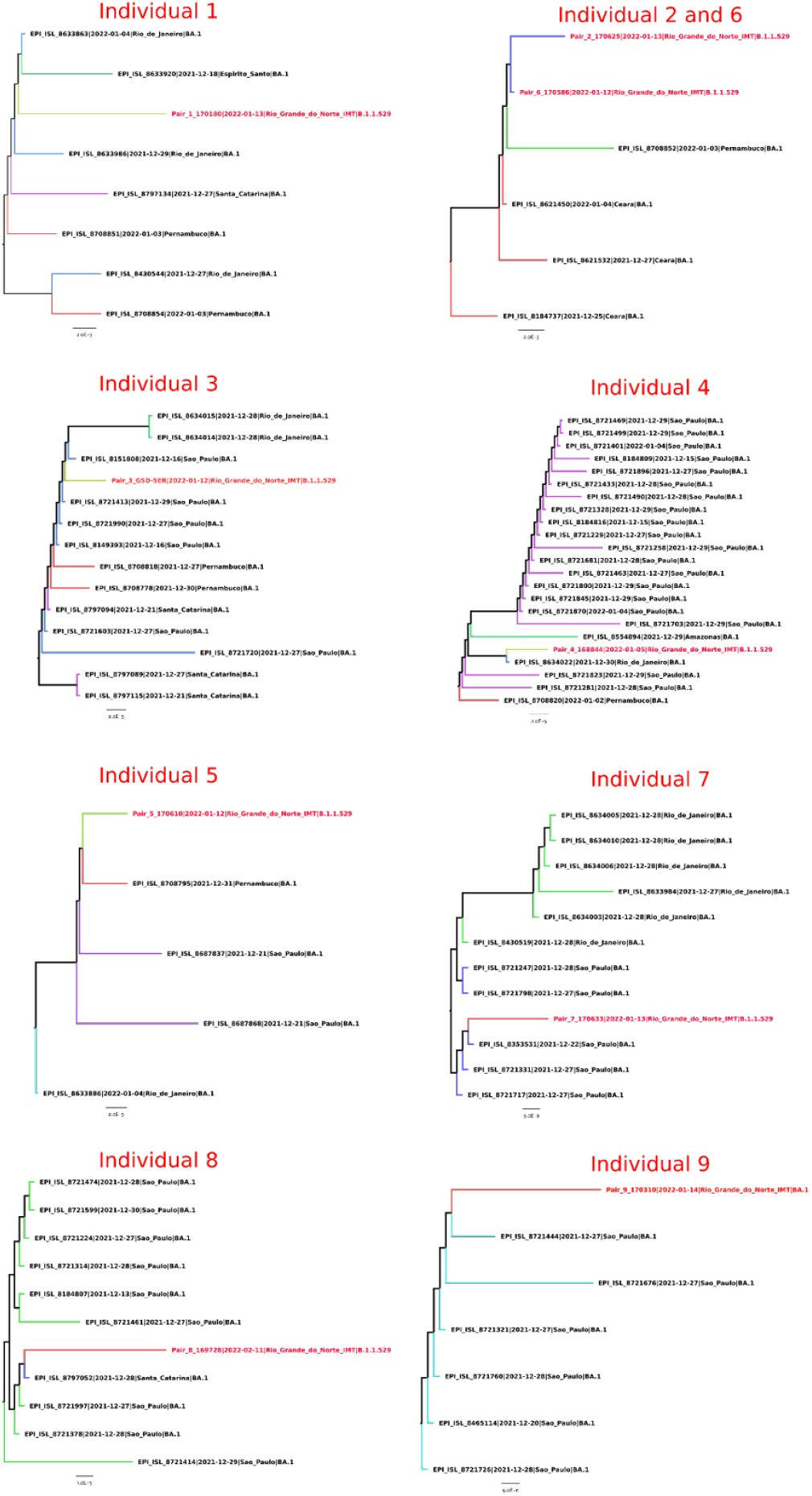
Subset of the Omicron (BA.1) ML phylogenetic tree indicating the closest isolates from different areas of Brazil. The Omicron genomes sequenced here are indicated with the red tips.

**Supplementary Figure S3.**
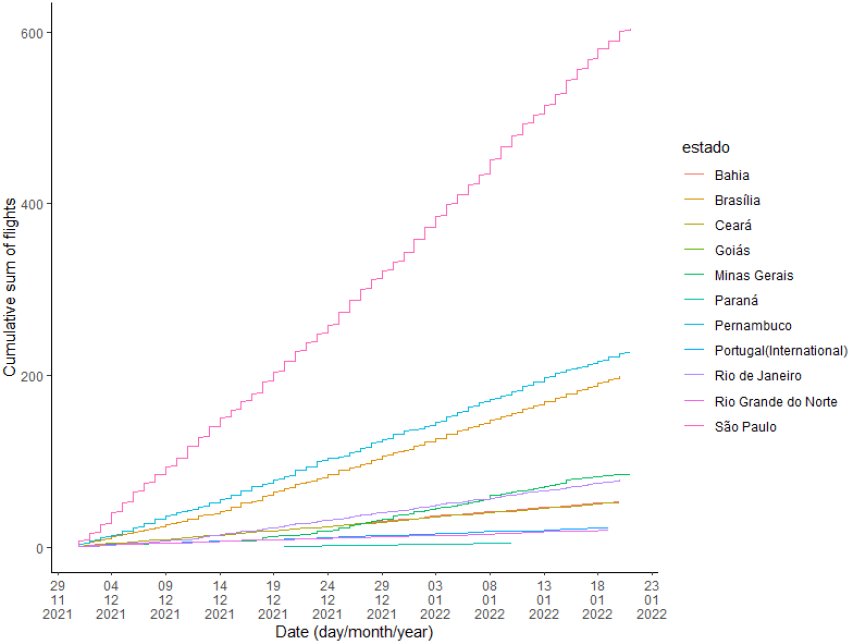
Cumulative sum of flights according to the Brazilian federative unit or Country of origin. From December 1, 2021 to December 31, 2021 the data was obtained from National Civil Aviation Agency (ANAC) - Consultation of past flights (https://sas.anac.gov.br/sas/bav/view/frmConsultaVRA, accessed on January 24, 2022). Data from January 1 2022 to January 20, 2022 the data was obtained from ANAC - Consultation of planned flights (https://siros.anac.gov.br/SIROS/(S(4gkz4ekn0yp1rg5lv3gvrq41))/view/registro/frmConsultaVoos, January 24, 2022). The order from the Brazilian federative unit with the most flight to Natal (from December 01, 2021 to January 20, 2022) from, to the lesser one, is: São Paulo (604 flights); Pernambuco (227 flights); Brasília (199 flights); Minas Gerais (85 flights); RIo de Janeiro (78 flights); Bahia (53 flights); Ceará (52 flights); Rio Grande do Norte (20 flights); Paraná (4 flights); Goiás (1 flight); and also Portugal with 22 international flights.

